# EFFECT OF TOBACCO CHEWING ON METABOLIC RISK FACTORS IN NORTH INDIAN ADULT MALES

**DOI:** 10.1101/2024.07.26.24311052

**Authors:** Abhishek Gupta, Arun Kumar Singh, Vani Gupta

## Abstract

Tobacco consumption remains one of the leading preventable causes of morbidity and mortality worldwide. Tobacco use is a major public health concern despite widespread public health efforts and strict regulations. Within the framework of the NCEP ATP-III criteria, the goal of the study was to investigate the effects of tobacco consumption on metabolic risk factors in young adult males. This case-control study comprised of 50 male participants aged 18 to 30 years; 25 male tobacco chewers were assigned to the study group, and 25 males who did not chew tobacco (non-tobacco chewers) were assigned to the control group. Anthropometric and metabolic risk factors that were compared between the study and control groups included waist circumference (WC), blood glucose, serum level of triglycerides (TG) and total cholesterol (TC), Insulin resistance (IR) and the homeostatic model assessment (HOMA) index, which is a measure of IR. The results of the study demonstrated that compared to non-tobacco chewers, tobacco chewers had significantly higher glucose (p=0.026), TC (p=0.0013), insulin (p=0.015), and IR (p=0.027). However, there were no discernible variations in HDL, VLDL, and TG levels, or WC. Nine male tobacco chewers and eight male non-tobacco chewers from the respective groups exhibited metabolic risk based on the NCEP ATP-III criterion of three out of five risk factors. Thus, we draw the conclusion that while tobacco chewers’ lipid profiles stayed similar, their TC, hyperglycaemia, hyperinsulinemia, and HOMA Index increased, making them more susceptible to the onset of metabolic syndrome.

## Introduction

The burden of non-communicable diseases is greatly increased by tobacco use, a serious worldwide health concern. Tobacco use is still common and affects millions of people globally, despite several public health measures aiming at lowering its prevalence. Although, the harmful effects of smoking on cardiovascular and respiratory health are widely known, concerns regarding the effects of smoking and smokeless tobacco products on metabolic health are also becoming more prevalent.

Many tobacco chewers are dying from serious illnesses like heart and lung conditions, mouth cancer, and other conditions. Chewing tobacco is the most popular way to consume tobacco. According to research, chewing tobacco is a common habit among youths over 25 year of age. Two-thirds of men chew tobacco in India, according to National Family Health Survey (NHFS) conducted in 2015–16. State-specific rates of chewing tobacco ranged from 15-45% among men (**1**). It is more prevalent among rural, underprivileged, and less educated people.

Due to the fact that nicotine induces IR, those who use it have a higher risk of developing diabetes. Nicotine use is closely linked to the level of IR and the severity of the accompanying metabolic problems. According to several experimental findings, smoking tobacco greatly reduces one’s ability to tolerate glucose, and smokers also have higher insulin levels (**2-4**). Today, a sizable portion of the global population is affected by the rise in nicotine consumption. It has a high risk of getting a number of diseases.

Adolescent metabolic syndrome and nicotine use have been found to be strongly correlated (**4**). IR increases the risk of developing diabetes mellitus and cardiovascular disease. Numerous prospective investigations have clearly demonstrated the link between IR and coronary artery disease. Its link to metabolic syndrome could help to explain some of the atherogenic effects of smoking. It has been discovered that smokers have a higher risk of developing diabetes than non-smokers. Furthermore, the relative risk is twice among middle-aged smokers compared to non-smokers after adjusting for age and BMI (**5**). Long-term use of smokeless tobacco is linked to a reduced risk of cardiovascular disease than smoking, while the risk for snuff users is higher than for non-smokers (**6**) and there is a notable increase in hypertension when compared to both smokers and non-smokers (**7**). However, there is a lack of research on the metabolic effects of smoking tobacco.

The amount of long-term nicotine usage measured by the plasma nicotine level was found to be strongly and negatively correlated with the degree of insulin sensitivity. There aren’t many studies on the immediate effects of smoking cessation on glucose metabolism. Using the euglycemic hyperinsulinemic clamp approach, ***Epifano et al***. demonstrated the effects of nicotine patches in type II diabetes patients and discovered a comparable outcome to that of smoking, namely decreased glucose utilization & storage, albeit to a lesser extent (**8**).

A straightforward recommendation for the routine identification of metabolic syndrome was made in the National Cholesterol Education Programme Adult Treatment Panel–III (NCEP ATP-III) report (**9**). This recommendation states that a person can be diagnosed with metabolic syndrome if they exhibit three of the following five characteristics: 1. Increased waist circumference (≥102 cm in man & ≥88cm in woman), 2. Elevated triglyceride (≥ 150mg/dl), 3. Reduced HDL cholesterol (<40mg/dl in man & < 50mg/dl in woman), 4. Elevated BP (<130/85 mm/Hg) or on treatment for hypertension, and 5. Elevated glucose (<100mg/dl). Consequently, a large body of research has demonstrated a causal link between IR and nicotine intake from smoking. Nonetheless, there aren’t many studies on tobacco use among non-smokers (tobacco chewers).

Therefore, the present study assessed the effects of short-term tobacco chewing (Gutka & Pan Masala) on metabolic risk factors using NCEP ATP-III in young, healthy male smokers who have not smoked but had chewed tobacco for at least five years and up to ten years. Males who were not smokers and who belonged to the same family (ideally a sibling or first-degree relative) and who were physically active were recruited based on their age, BMI, and diet. Thus, we can lessen the impact of additional confounding factors. Study examined the IR in tobacco chewers and non-tobacco chewers using the HOMA index with NCEP ATP-III criteria 2001, to determine the presence of metabolic risk factors.

## Methods

### Selection of study subjects and diagnostic criteria

A case-control study was conducted here. There were fifty participants in the study, all between the ages of 18 and 30. Twenty-five of these were male tobacco chewers (diagnosed using NCEP ATP-III criteria, 2001) who were recruited from Lucknow city; the other twenty-five males who were not tobacco chewers were picked as the control group based on age and BMI matching. While metabolic or systemic disease, alcohol addiction, smoking, BMI less than 21 or more than 25, WHR more than 0.90, and age less than 18 and more than 30 years were excluded, all of the male participants in the study appeared to be in good health. They also had a BMI between 21 and 25 and a WHR of less than 0.90. The subjects gave their informed consent to participate in the study, and King George’s Medical University in Lucknow’s ethics committee approved it.

### Estimation of biochemical parameters

Within three months, all the males were examined. Male’s waist to hip ratios of greater than 0.90 and body mass index between 21 and 25 were measured. Every individual had 5 ml of blood sample drawn after overnight fasting, and the plasma/serum was separated out and stored in adequate vials and temperature for serum lipid profile, insulin, and glucose levels. With the GOD-POD method (Randox Laboratories Ltd., Antrim, UK), plasma glucose was estimated (**10**). By using an enzymatic approach (Randox Laboratories Ltd., Antrim, UK) (**11**) and an immuno-radiometric test method (Immunotech Radiova, Prague), serum insulin was measured (**12**). The IR was determined using the Homeostatic Model Assessment Index (HOMA Index) (**13**).

HOMA Index = (Insulin in *μ* unit / ml) / 22.5 X e^- (in fasting glucose m.mole / L)^

A laboratory diagnosis of IR is made when the HOMA Index value equals or exceeds 3.8 in people who did not meet the clinical and biological criteria for the condition. Cancer development is more likely in subjects with a HOMA Index greater than 3.8.

### Statistical analysis

All data was statistically analysed by using ‘unpaired student t test’ and multiple logistic regression and Fisher-exact test. Whenever necessary, χ2 analysis was carried out. Accordingly, odds ratios and 95% confidence intervals are provided; statistical significance was defined as p values less than 0.05. We used GraphPad Prism 7 (version 7.0), a commercially available program, for statistical analysis.

## Result

Tobacco chewers and non-tobacco chewers were compared with respect to the existence of various risk factors, such as WC, blood glucose, serum TC, TG, HDL, VLDL, insulin and IR are shown **in Table 1**. Between male tobacco chewers and non-tobacco chewers, there was no statistically significant difference in WC and in the levels of HDL, VLDL, and TG. Compared to non-tobacco chewers, tobacco chewers had significantly greater blood glucose, serum TC, insulin, and HOMA-IR (**Table 1**).

**Table 1:**
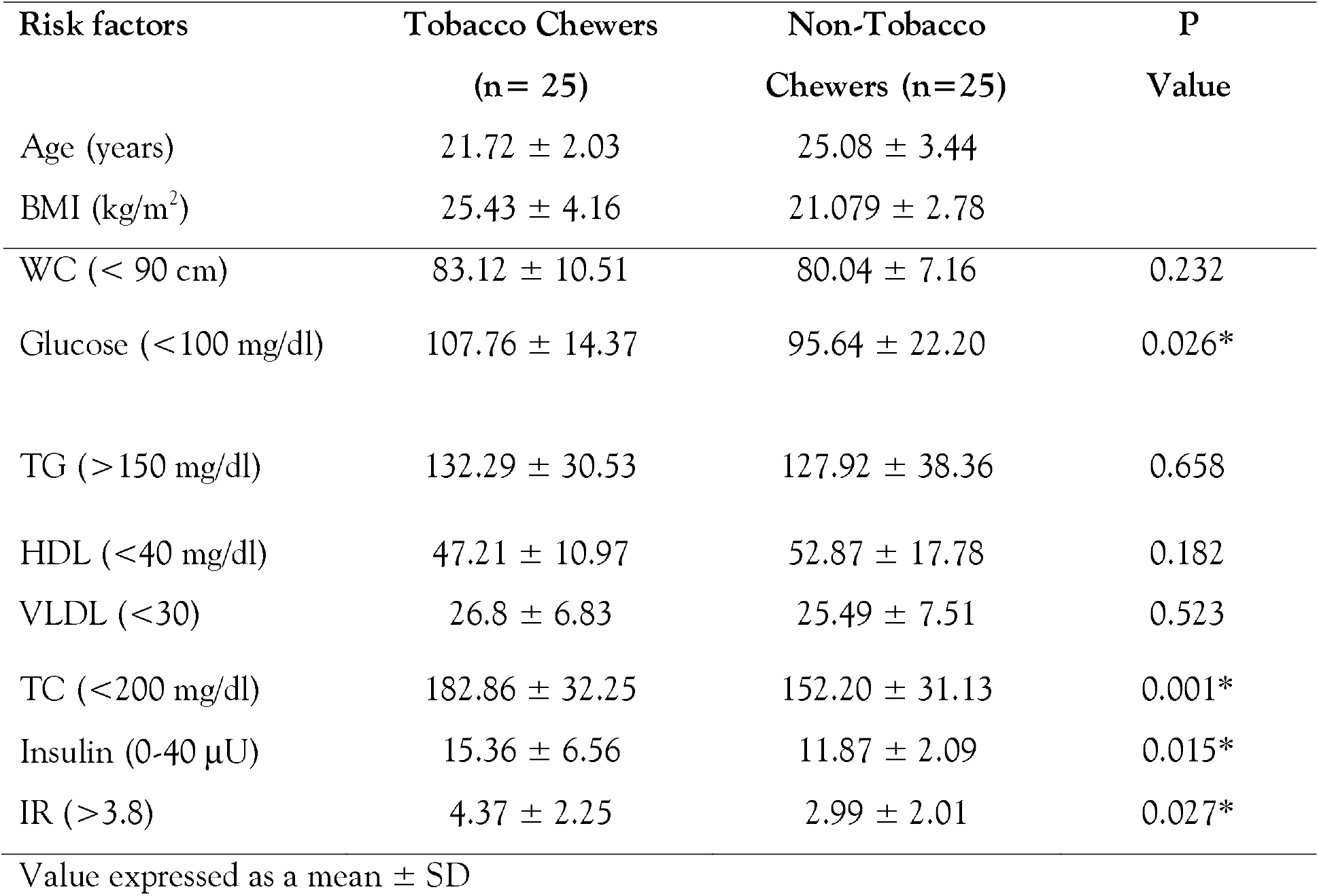
Comparison of risk factors between tobacco chewers and non-tobacco chewers according to NCEP ATP-III, 2001 criteria.

| Risk factors | Tobacco Chewers<br>(n= 25) | Non-Tobacco<br>Chewers (n=25) | P<br>Value |
| --- | --- | --- | --- |
| Age (years) | 21.72 ± 2.03 | 25.08 ± 3.44 | |
| BMI (kg/m<sup>2</sup>) | 25.43 ± 4.16 | 21.079 ± 2.78 | |
| WC (< 90 cm) | 83.12 ± 10.51 | 80.04 ± 7.16 | 0.232 |
| Glucose (<100 mg/dl) | 107.76 ± 14.37 | 95.64 ± 22.20 | 0.026* |
| TG (> 150 mg/dl) | 132.29 ± 30.53 | 127.92 ± 38.36 | 0.658 |
| HDL (<40 mg/dl) | 47.21 ± 10.97 | 52.87 ± 17.78 | 0.182 |
| VLDL (<30) | 26.8 ± 6.83 | 25.49 ± 7.51 | 0.523 |
| TC (<200 mg/dl) | 182.86 ± 32.25 | 152.20 ± 31.13 | 0.001* |
| Insulin (0-40 μU) | 15.36 ± 6.56 | 11.87 ± 2.09 | 0.015* |
| IR (>3.8) | 4.37 ± 2.25 | 2.99 ± 2.01 | 0.027* |

Value expressed as a mean ± SD

Tobacco chewers showed significantly higher levels of fasting blood glucose, insulin, TC, and HOMA-IR (all p <0.05) compared to non-tobacco chewers (p >0.05). While a statistically significant difference in WC, serum HDL, TG, and VLDL levels were not identified between tobacco chewers and non-tobacco chewers, a significantly larger number of participants were reported to have HOMA-IR >3.8 in tobacco chewers are given in **Table 1**.

BMI, body mass index; WC, waist circumference; TG, triglyceride; HDL, high density lipoprotein; VLDL, very low-density lipoprotein; TC, total cholesterol; insulin resistance, IR

According to NCEP-ATP-III criteria of three out of five risk factors, nine tobacco chewers and eight non-tobacco chewers males from respective groups had metabolic risk. Fifteen tobacco chewers and six non-tobacco chewers from respective groups had HOMA Index >3.8 are shown in Table 2.

**Table 2:**
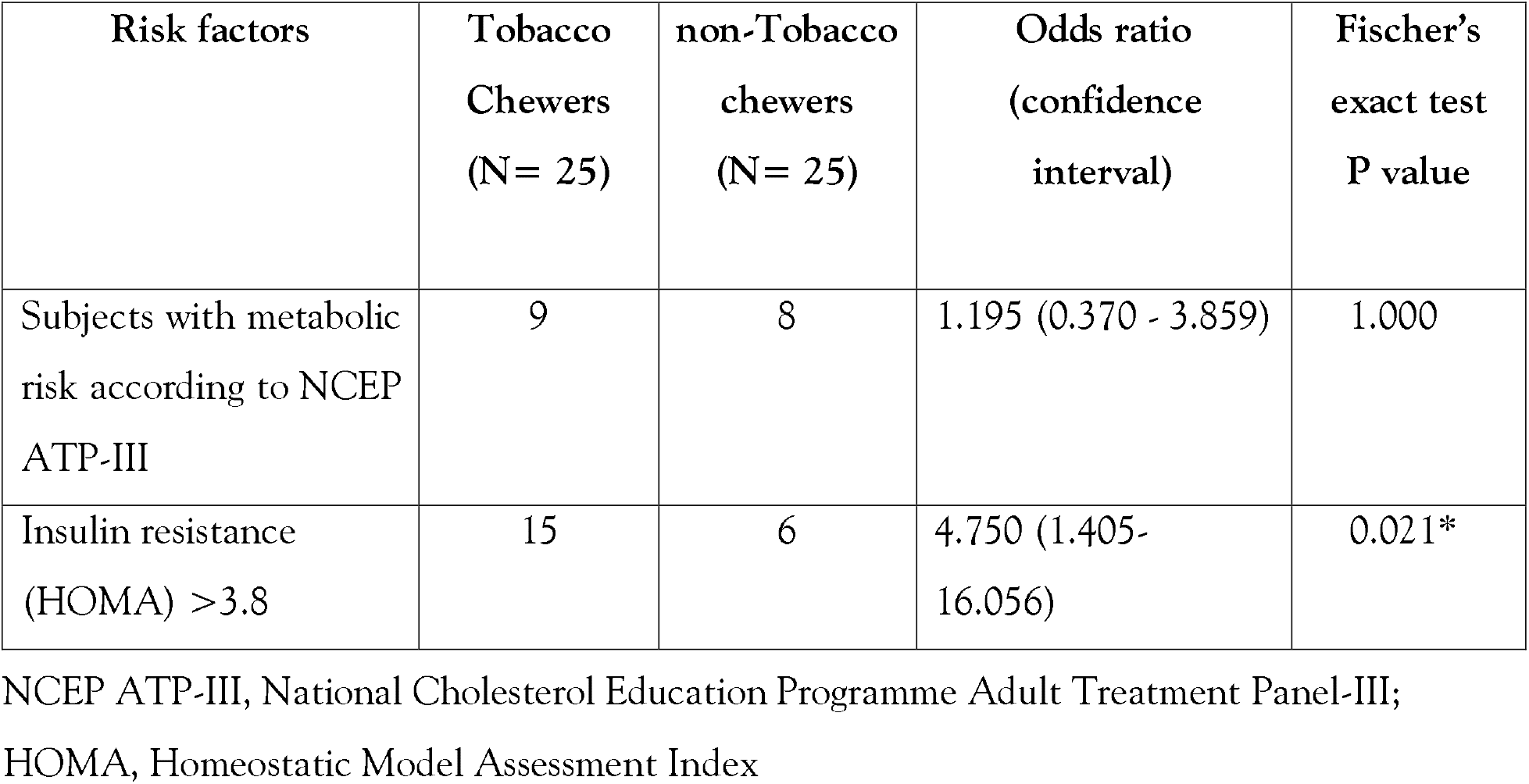
Comparison of exposure to risk factors between Tobacco and non-Tobacco chewers.

| **Risk factors** | **Tobacco Chewers (N= 25)** | **non-Tobacco chewers (N= 25)** | **Odds ratio (confidence interval)** | **Fischer's exact test P value** |
| --- | --- | --- | --- | --- |
| Subjects with metabolic risk according to NCEP ATP-III | 9 | 8 | 1.195 (0.370 - 3.859) | 1.000 |
| Insulin resistance (HOMA) >3.8 | 15 | 6 | 4.750 (1.405-16.056) | 0.021* |

NCEP ATP-III, National Cholesterol Education Programme Adult Treatment Panel-III; HOMA, Homeostatic Model Assessment Index

## Discussion

Globally, there is a wealth of research indicating that the tobacco pandemic is mostly caused by young tobacco users. It is also linked to a complex etiology that youth tobacco use. In contrast to the Western world, where it has been extensively studied, emerging nations like India have given this less consideration. In India, there are a plethora of handmade and commercial smokeless tobacco products accessible. Chewing tobacco are more common than snuff use. In this case control study, two male groups—those who chewed tobacco and those who did not—were similar in terms of sex, age, BMI, level of physical activity, diet, and even genetic makeup (since we divided the groups into those who chewed tobacco for five to ten years and those who did not). The subjects appeared to be in good health, with a BMI ranging from 21 to 25, a male adult WHR of less than 0.90, and an age group of 18 to 30 years.

The impact of non-smoking tobacco usage on risk factors was investigated in the study using the NCEP ATP-III criteria, 2001. On the other hand, because nicotine produces IR, previous research has demonstrated that those who use nicotine have an increased risk of developing diabetes mellitus (**14,15**). In comparison to non-tobacco chewers, findings revealed that tobacco chewers had significantly higher fasting serum insulin, TC, blood glucose, and HOMA Index (**Table 1**). Nevertheless, IR>3.8 was present in 15 tobacco chewers and 6 non-tobacco chewers. However, chewing tobacco results in elevated IR, as seen by the much higher HOMA Index value among tobacco chewers compared to non-tobacco chewers. The rise in IR observed in tobacco chewers may be due to a mechanism that affects multiple metabolic pathways simultaneously or to an early step in the action of insulin, such as glucose transport, signal transduction, and/or glucose phosphorylation. When nicotine interacts with insulin receptors and causes post-receptive events, it can have a direct or indirect impact. Chewing tobacco increases catecholamine release, which may lead to a decrease in insulin binding sites and a reduction in the production of glucose transporters (**16**). Chewing tobacco may potentially have further unintended consequences for the way insulin works. For instance, high free fatty acid levels can hinder the absorption of glucose mediated by insulin.

Compared to non-tobacco chewers, tobacco chewers in the study had higher fasting insulin levels. Regarding the variables known to affect insulin action and glucose intolerance, the two subject groups were similar. It would therefore seem reasonable to hypothesize that the difference in fasting insulin. The differences we saw between the two groups were primarily due to tobacco chewing, and they might have been brought on by the nicotine or other chemicals that tobacco chewers directly consumed. The study results showing that chewing tobacco can temporarily reduce the effectiveness of insulin. These current results are consistent with previously published research that showed chewing tobacco increases insulin sensitivity and hyperinsulinemia, which raises the chance of developing diabetes mellitus (**14,15,17,18**). A few research, however, reported the opposite results (**19,20**).

The study also examined the degree to which the fasting serum cholesterol levels of tobacco chewers and non-tobacco chewers differed (**Table 1**). We may not have identified a significant statistical difference in the fasting lipid profile between tobacco chewers and non-tobacco chewers since our sample includes young male subjects between the ages of 18 and 30, as well as college students. Those who have chewed tobacco for five to ten years are among the registered participants in the study. The lipid profile of tobacco chewers did not alter; however, they did have elevated TC, blood glucose, hyperinsulinemia, and HOMA Index. Thus, the study concluded that tobacco consumption is more prone for the development of metabolic syndrome.

## Data Availability

All data produced in the present work are contained in the manuscript

## Acknowledgement

We would like to thank faculty and staff of the Physiology and Medicine Department, KGMU, Lucknow for providing tobacco chewers subjects.

